# Prevalence of orthostatic intolerance in Long Covid clinic patients: A multicentre observational study

**DOI:** 10.1101/2023.12.18.23299958

**Authors:** Cassie Lee, Darren C Greenwood, Harsha Master, Kumaran Balasundaram, Paul Williams, Janet T. Scott, Conor Wood, Rowena Cooper, Julie L. Darbyshire, Ana Espinosa Gonzalez, Helen E. Davies, Thomas Osborne, Joanna Corrado, Nafi Iftekhar, Natalie Rogers, Brendan Delaney, Trish Greenhalgh, Manoj Sivan, the LOCOMOTION Consortium

**Author notes:** **Corresponding Author** Cassie Lee, ICRRU, Mint Wing, St. Mary’s Hospital, Imperial College HealthCare NHS Trust, Praed Street, W2 1NY. **Details of roles and contributors.** Name of guarantor – Dr Manoj Sivan Miss Cassie Lee, Clinical Research fellow: Investigation, Methodology, Visulisation, Writing - original draft, Writing - review and editing. Dr Darren C Greenwood, Senior Lecturer in Biostatistics: Data curation, Formal Analysis, Visulisation, Writing - original draft, Writing - review and editing. Dr Harsha Master, GP Lead Covid Rehabilitation Service: Investigation, Visulisation, Writing - original draft, Writing - review and editing. Dr Kumaran Balasundaram, Clinical Research Fellow: Investigation, Writing - original draft, Writing - review and editing. Dr Paul Williams, Clinical Research Fellow: Investigation, Writing - original draft, Writing - review and editing. Dr Janet T. Scott, Consultant in Infectious Disease & Research Medicine: Methodology, Writing - review and editing Mr Conor Wood, Research Assistant: Investigation, Writing - review and editing. Dr Rowena Cooper, Specialty Research Doctor: Investigation, Writing - review and editing. Ms Julie L. Darbyshire, Senior Scientific Officer: Conceptulisation, Methodology, Writing - review and editing. Dr Ana Espinosa Gonzalez, Clinical Research Fellow, Methodology, Writing - original draft. Dr Helen E. Davies, Consultant in Respiratory Medicine: Methodology, Writing - review and editing. Mr Thomas Osborne, Research Project Manager: Project Administration, Writing - review and editing. Dr Joanna Corrado, Specialist Trainee in Rehabilitation Medicine: Investigation, Methodology. Dr Nafi Iftekhar, Medical student: Investigation, Methodology. Ms Natalie Rogers, Patient with lived experience & Member of the PAG: Conceptulisation, Writing - review and editing. Professor Brendan Delaney, Chair in Medical Informatics and Decision Making: Conceptulisation, Funding acquisition, Methodology, Writing - review and editing. Professor Trish Greenhalgh, Professor of Primary Care Health Sciences: Conceptulisation, Methodology, Visulisation, Writing - original draft, Writing - review and editing. Dr Manoj Sivan, Associate Clinical Professor and Honorary Consultant in Rehabilitation Medicine: Conceptulisation, Funding acquisition, Methodology, Visulisation, Writing - review and editing, Supervision. The Corresponding Author has the right to grant on behalf of all authors and does grant on behalf of all authors, a worldwide licence to the Publishers and its licensees in perpetuity, in all forms, formats and media (whether known now or created in the future), to i) publish, reproduce, distribute, display and store the Contribution, ii) translate the Contribution into other languages, create adaptations, reprints, include within collections and create summaries, extracts and/or, abstracts of the Contribution, iii) create any other derivative work(s) based on the Contribution, iv) to exploit all subsidiary rights in the Contribution, v) the inclusion of electronic links from the Contribution to third party material where-ever it may be located, and vi) licence any third party to do any or all of the above. **Transparency declaration** The lead author and the manuscript’s guarantor affirms that the manuscript is an honest, accurate, and transparent account of the study being reported; that no important aspects of the study have been omitted; and that any discrepancies from the study as planned (and, if relevant, registered) have been explained. **Statements and Declarations** This work is independent research funded by the National Institute for Health and Care Research (NIHR) (Long Covid grant, Ref: COV-LT2-0016). The views expressed in this publication are those of the authors and not necessarily those of NIHR or The Department of Health and Social Care.

## Abstract

**Purpose:** Orthostatic intolerance (OI), including postural orthostatic tachycardia syndrome (PoTS) and orthostatic hypotension (OH), are often reported in long covid, but published studies are small with inconsistent results. We sought to estimate the prevalence of objective OI in patients attending long covid clinics and healthy volunteers and associations with symptoms and comorbidities.

**Methods:** Participants were recruited from 8 UK long covid clinics, and healthy volunteers from general population. All undertook standardised National Aeronautics and Space Administration Lean Test (NLT). Participants’ history of typical OI symptoms (e.g. dizziness, palpitations) prior to and during the NLT were recorded.

**Results:** 277 long covid patients and 50 frequency-matched healthy volunteers were tested. Healthy volunteers had no history of OI symptoms or PoTS, 10% had asymptomatic OH. 130 (47%) long covid patients had previous history of OI symptoms and 144 (52%) developed symptoms during the NLT. 41 (15%) had an abnormal NLT, 20 (7%) met criteria for PoTS and 21 (8%) had OH. Of patients with an abnormal NLT, 45% had no prior symptoms of OI. Relaxing the diagnostic thresholds for PoTS from two consecutive to one reading, resulted in 11% of long covid participants meeting criteria for PoTS, but not in healthy volunteers.

**Conclusion:** More than half of long covid patients experienced OI symptoms during NLT and more than one in ten patients met the criteria for either PoTS or OH, half of whom did not report previous typical OI symptoms. We recommend all patients attending long covid clinics are offered an NLT and appropriate management commenced.

Trial registration numbers NCT05057260, ISRCTN15022307

## Introduction

The protracted form of covid-19 known as long covid (1–3) is frequently accompanied by orthostatic intolerance (OI) (4,5) that is, symptoms suggestive of an abnormal haemodynamic response to upright posture, such as dizziness, light-headedness, palpitations or tremulousness.(6) During objective testing, the blood pressure may fall significantly on standing, (orthostatic hypotension, OH) (7,8) *or* there may be a significant rise in heart rate on standing *without* a drop in blood pressure—a condition known as orthostatic tachycardia (OT), or (if accompanied by symptoms during testing) postural orthostatic tachycardia syndrome (PoTS).(6,9) The diagnosis of PoTS requires OH to be excluded, hence a patient may have either OH or PoTS but not both. Both OH and PoTS occur in several different conditions and are thought to be due to abnormal function of the autonomic nervous system.(6,10,11) OH is typically found in cases such as older people, people with diabetes, and people on diuretics.(7,8) PoTS tends to occur more in younger people, and can incur diagnostic delays, with often-marked functional impairment and psychological impact.(12,13) Some studies have suggested a link with PoTS and myalgic encephalomyelitis / chronic fatigue syndrome.(14–18) Orthostatic abnormalities including OH, OT and PoTS have also been identified as a sequel of long covid,(5,19–24) which can have a significant impact on day-to-day functioning.(25) long covid is associated with many and diverse abnormalities in the neurological and cardiovascular systems, including abnormal autonomic response (‘dysautonomia’).(10,26,27) Symptoms of long covid can be complex and prolonged,(28) many of which overlap with those of OI, and include dizziness, palpitations, tightness or pain in the chest, and tingling in the extremities on standing.(4,29) The presentation of non-specific OI symptoms, such as generalized tiredness and cognitive impairment,(6) can render a diagnosis of OH or PoTS difficult to detect.(29,30) This is further compounded given the lack of association in long covid with specific symptoms and those associated with typical OI,(30–32) There have been several previous studies on the prevalence of OH and PoTS in long covid patients,(24) but they are small or single-clinic studies,(5,30) and samples were sometimes skewed. Prevalence of OI is varied and ranges from 2% to 33% (20,24) with a reported incidence of 71% obtained from a sample of 29 patients with OI referred for tilt table testing (a specialist test not available in standard clinics.(5)

This study was designed to answer three important questions about OI in long covid using a large sample drawn from multiple clinics. First, what is the overall prevalence of OH and PoTS in an unselected clinic population using a standard objective test? Second, can people with OH and PoTS be identified purely by testing those who reported typical OI symptoms such as palpitations or dizziness on standing, or is formal testing of everyone necessary? And third, what associations are there between long covid’s features, comorbidities and either OH or PoTS?

## Method

### The LOCOMOTION study

The work reported here was part of LOCOMOTION (LOng COvid Multidisciplinary consortium Optimising Treatments and services across the NHS), a 30-month multi-site case study of ten long covid clinics (of which eight participated in this sub-study), beginning in 2021, which seeks to optimise long covid care across the clinics. Each clinic offers multidisciplinary care to patients referred from primary care (and, in some cases, self-referred). A study protocol for LOCOMOTION, with details of management, governance and patient involvement has been published (33). Ethics approval was granted by Yorkshire & The Humber— Bradford Leeds Research Ethics Committee (REC; ref: 21/YH/0276) and subsequent amendments.

### Patient involvement

One work package in LOCOMOTION involves a quality improvement collaborative, in which front-line clinicians from all ten clinics and patient advisors identify priority areas for improvement and follow a cycle of measuring current practice, implementing change, and re-measuring practice [paper submitted]. The study concept and question of whether PoTS was being diagnosed and adequately managed in these clinics was the leading priority for our patient advisers and among the top questions raised by clinicians.

### Sampling and data sources

Based on previous literature (18,34) we estimated that 250 patients would provide sufficient precision to estimate 30% positive NLT to within ±6%. With 50 healthy volunteers this would also provide 90% power to detect a 20% difference between groups in positive NLTs (30% vs 10%) at p<0.05.

Data collection occurred from May to October 2023 in eight clinics across the UK. Consecutive patients attending participating clinics were invited to join the study. Inclusion criteria were age over 18, able and willing to consent, either a long covid clinic patient or a healthy volunteer, and (if self-testing at home) in possession of a blood pressure machine. The healthy volunteers were age-sex frequency-matched to the long covid group using ten-year age-bands.

Exclusion criteria were inability to give informed consent or comply with test instructions, if the clinical team considered the test unsuitable or unsafe (e.g., if the patient could not stand unaided), and any coexisting condition that could interfere with autonomic or haemodynamic function (e.g., pregnancy). Verbal informed consent to perform the NLT were obtained from all participants. Written consent were obtained by the healthy volunteers, this was considered unnecessary for the long covid patient since this intervention was already part of standard clinical assessment in many clinics and its introduction had been justified as a quality improvement measure.

### Orthostatic Intolerance (OI) symptoms

Prior to having the test, participants were asked about their long covid symptoms specifically to detail typical symptoms suggestive of OI (6): dizziness, palpitations, chest pain or discomfort, tremulousness.

### The NASA Lean Test

The National Aeronautics and Space Administration Lean Test (NLT) was used as per published instructions. (35) Participants were offered the test as part of a clinic assessment or given the option of self-administering at home for the few sites that offered this.

In the NLT, the participant first lies quietly for two to five minutes, then two supine readings (pulse and blood pressure) were taken one minute apart. The participant then stands up slowly with the shoulders leaning against a wall (for support), with feet about 15 cm from the wall. Six further readings were taken at one-two min intervals. [See supplementary material, Lean test protocol, for more details]

The NLT was terminated prematurely if participants’ symptoms were such that they were unable to complete the full 10 minutes or if the clinician was concerned. If the NLT was stopped early, available data were analysed.(30) Regular meetings were held to ensure consistency in practice across participating sites.

Table 1 shows the criteria we used for diagnosing OH, PoTS, OI, symptomatic NLT, and a positive NLT.

**Table 1.** Definitions used to diagnose or exclude OH and PoTS in the NASA Lean Test (NLT)

|  |  |
| --- | --- |
| Orthostatic Intolerance (OI) | Dizziness/light-headedness, palpitations, chest pain, or tremulousness which get worse when sitting or standing and improves or resolves when lying down |
| Orthostatic Hypotension (OH) | Fall in systolic blood pressure within the first three readings of at least 20 mmHg, or fall in diastolic blood pressure of at least 10 mmHg with or without acute symptoms during test |
| Orthostatic Tachycardia (OT) | Increase in HR above baseline of at least 30 bpm, sustained for two consecutive readings, after excluding OH |
| Postural Orthostatic Tachycardia Syndrome (PoTS) | OT plus symptoms of OI during the NLT |
| Symptomatic NLT | Any symptoms reported during the NLT, including baseline symptoms that were exacerbated by standing |
| Positive NLT | Patients meeting the criteria for PoTS or OH |
| Narrow pulse pressure (PP)* | Difference between systolic and diastolic blood pressure which is less than 25% of the systolic blood pressure |

\*(38),(34)

### Objectives

We aimed to estimate (1) the prevalence of subjective OI (symptoms of orthostatic intolerance) and objective OI (OH and PoTS) in healthy people and those living with long covid; (2) the association between patient characteristics and prevalence of objective OI, OH, and PoTS; (3) the prevalence of OH and PoTS in people with a history of OI symptoms compared to those without a history; (4) the impact of relaxing the standard thresholds for the diagnosis of PoTS.

### Data management

Anonymised data were shared with the host site, University of Leeds, for data analysis, adhering with sites’ local governance procedures and in compliance with the ethics approvals.

### Statistical analysis

All statistical analyses were conducted in Stata version 18. [StataCorp. 2023. Stata: Release 18. Statistical Software. College Station, TX: StataCorp LLC.] 95% confidence intervals were calculated for proportions of participants with sustained increased in HR, OI, OH, narrow PP, and PoTS during their NLT. Where there were no participants in a category, one-sided exact 97.5% confidence intervals were calculated. Comparisons between two proportions used Fisher’s exact test, with exact 95% confidence intervals.

To identify predictors of PoTS or OH in long covid patients, logistic regression was used to quantify associations between patient characteristics (age, sex, body mass index, ethnicity), pre-existing comorbidities, and prior symptoms (see list above) and confirmation of PoTS following an NLT. Models were calculated both unadjusted and adjusted for age and sex. Categories with fewer than 10 participants were merged or excluded. Where an NLT was terminated early, the last measurements were carried forward.

### Sensitivity analyses

To understand whether long covid patients were experiencing symptoms at lower thresholds than the formal definition of PoTS, we explored the robustness of our results when applying different definitions. We evaluated sensitivity to how long an increased heart rate needed to be sustained for (over two minutes, any two consecutive time points, any two time points, or any one time point).

## Results

### Participants

Across eight clinics, 277 patients and 50 healthy volunteers frequency-matched on age and sex were recruited. Participant characteristics are shown in Table 2. The demographic characteristics of long covid patients recruited to this study broadly reflected those of Long Covid clinic populations.(36) (Table 2) Healthy volunteers were matched on age and sex but had lower mean body mass index (BMI) and fewer comorbidities than long covid patients (Table 2).

**Table 2.** Characteristics of participants.

|  | long covid patients<br>(n=277) | Healthy volunteers<br>(n=50) |
| --- | --- | --- |
| Mean age (years) (SD) | 48 (13) | 48 (16) |
| Sex: |  |  |
| *Female (%)* | 173 (62%) | 32 (64%) |
| *Male (%)* | 104 (38%) | 18 (36%) |
| Mean body mass index (kg/m<sup>2</sup>) (SD) | 29 (7) | 25 (5) |
| Ethnicity: |  |  |
| *White (%)* | 219 (79%) | 31 (62%) |
| *Black (%)* | 7 (3%) | 0 (0%) |
| *Asian (%)* | 23 (8%) | 3 (6%) |
| *Mixed/other (%)* | 16 (6%) | 14 (28%) |
| *Not recorded (%)* | 12 (4%) | 2 (4%) |
| Known medical conditions: |  |  |
| *long covid* | 277 (100%) | 0 |
| *Allergies or autoimmune conditions (%)* | 44 (16%) | 6 (12%) |
| *Other respiratory conditions (%)* | 1 (<1%) | 0 (0%) |
| *Other inflammatory conditions (%)* | 12 (4%) | 2 (3%) |
| *Hypertension (%)* | 28 (10%) | 4 (6%) |
| *Hypotension (%)* | 0 (0%) | 0 (0%) |
| *Other heart conditions (%)* | 22 (8%) | 2 (4%) |
| *Type 2 diabetes mellitus (%)* | 16 (6%) | 1 (2%) |
| *Mental health condition (%)* | 43 (16%) | 0 (0%) |
| Median duration of long Covid (IQR)<br>(months) | 18 (14 to 28) | - |
| Admitted to hospital with initial SARS-CoV-2 infection (%) * | 16 (9%) | - |
| Admitted to intensive care with initial<br>SARS-CoV-2 infection (%) * | 4 (2%) | - |

\* Where numbers do not sum to totals, this is because of incomplete data

### Prevalence of OI, OH, and PoTS in healthy volunteers

None of the healthy volunteers had symptoms of OI prior to testing or had to terminate the NLT early because of symptoms. None of these participants had orthostatic tachycardia 0/50 (0%) (95% CI: 0% to 7%) or met the criteria for PoTS (Table 3). Five (10%, 95% CI: 3% to 22%) met the haemodynamic criteria for OH (Table 3), four were identified within two minutes of standing and one at minute four. All healthy volunteers had no previous history suggestive of OI and were asymptomatic during the test.

**Table 3.** Proportion of long covid patients (both with and without a previous history of OI) and health volunteers meeting criteria for PoTS and OH.

|  | long covid patients (n=277) |  |  |  |
| --- | --- | --- | --- | --- |
|  | With<br>previous<br>history of OI<br>(n=130) | No previous<br>history of OI<br>(n=147) | All long covid<br>patients<br>(n=277) | Healthy volunteers<br>(n=50) |
| Met criteria for PoTS (%)<br>(95% CI) | 14 (11%)<br>(6%, 17%) | 6 (4%)<br>(2%, 9%) | 20 (7%)<br>(4%, 11%) | 0 (0%)<br>(0%, 7%) |
| Met criteria for OH with or<br>without symptoms during<br>Lean Test (%)<br>(95% CI) | 7 (5%)<br>(2%, 11%) | 14 (10%)<br>(5%, 15%) | 21 (8%)<br>(5%, 11%) | 5 (10%)<br>(3%, 22%) |

### Prevalence of OI, OH, and PoTS in long covid clinic patients

#### Orthostatic Intolerance

Of the 277 patients recruited from long covid clinics, 130 (47%) had a previous history of symptoms suggestive of OI (dizziness, palpitations, chest pain or tremors) prior to their NLT. A total of 144 (52%) were symptomatic during the NLT, with a range of acute symptoms reported (most commonly dizziness but also palpitations, chest pain or tremors, fatigue, muscle or joint pain, cognitive dysfunction, headache, visual disturbances, breathlessness, tingling or numbness in the skin, sweating or clamminess) (Table 4). In the NLT, 20 (7%, 95% CI: 4 to 11%) long covid patients met the criteria for PoTS and 21 (8%, 5 to 11%) met the criteria for OH (Table 3), 18 reached the threshold for OH within the first two minutes of standing and three by minute four. A total of 46 (17%) patients stopped the test early because of excessive symptoms. Of these, just six met the criteria for PoTS and seven met the criteria for OH (see supplementary Table 1 for details of tests ended prematurely).

**Table 4.** Proportion of acute test symptoms of dysautonomia on standing presenting in long covid patients and healthy controls during Lean Test, with 95% confidence intervals.

| Symptom | In all long covid patients<br>(n=277) | In those with prior history of OI symptoms<br>(n=130) | In those meeting criteria for PoTS<br>(n=20) | In those meeting criteria for OH<sup>†</sup><br>(n=21) | In healthy volunteers<br>(n=50) |
| --- | --- | --- | --- | --- | --- |
| *All symptoms:* |  |  |  |  |  |
| Any symptoms during Lean Test | 144 (52%)<br>(46%, 58%) | 84 (65%)<br>(56%, 73%) | 20 (100%)<br>(83%, 100%)* | 11 (52%)<br>(30%, 74%) | 0 (0%)<br>(0%, 7%) |
| *Symptoms of orthostatic intolerance:* |  |  |  |  |  |
| Dizziness / light-headedness | 92 (33%)<br>(28%, 39%) | 67 (52%)<br>(43%, 60%) | 13 (65%)<br>(41%, 85%) | 5 (24%)<br>(8%, 47%) | 0 (0%)<br>(0%, 7%) |
| Palpitations | 15 (5%)<br>(3%, 9%) | 13 (10%)<br>(5%, 16%) | 2 (10%)<br>(1%, 32%) | 0 (0%)<br>(0%, 16%) | 0 (0%)<br>(0%, 7%) |
| Chest pain / discomfort | 12 (4%)<br>(2%, 7%) | 10 (8%)<br>(4%, 14%) | 3 (15%)<br>(3%, 38%) | 1 (5%)<br>(0%, 24%) | 0 (0%)<br>(0%, 7%) |
| Tremors | 2 (1%)<br>(0%, 3%) | 2 (2%)<br>(0%, 5%) | 0 (0%)<br>(0%, 17%) | 0 (0%)<br>(0%, 16%) | 0 (0%)<br>(0%, 7%) |
| *Other symptoms of dysautonomia:* |  |  |  |  |  |
| Tingling / numbness in skin | 34 (12%)<br>(9%, 17%) | 18 (14%)<br>(8%, 21%) | 6 (30%)<br>(12%, 54%) | 2 (10%)<br>(1%, 30%) | 0 (0%)<br>(0%, 7%) |
| Fatigue | 26 (9%)<br>(6%, 13%) | 6 (5%)<br>(2%, 10%) | 1 (5%)<br>(0%, 25%) | 3 (14%)<br>(3%, 36%) | 0 (0%)<br>(0%, 7%) |
| Sweating / clammy / hot | 19 (7%)<br>(4%, 11%) | 10 (8%)<br>(4%, 14%) | 2 (10%)<br>(1%, 32%) | 3 (14%)<br>(3%, 36%) | 0 (0%)<br>(0%, 7%) |
| Breathlessness | 15 (5%)<br>(3%, 9%) | 7 (5%)<br>(2%, 11%) | 3 (15%)<br>(3%, 38%) | 0 (0%)<br>(0%, 16%) | 0 (0%)<br>(0%, 7%) |
| Headache | 11 (4%)<br>(2%, 7%) | 6 (5%)<br>(2%, 10%) | 5 (25%)<br>(9%, 49%) | 1 (5%)<br>(0%, 24%) | 0 (0%)<br>(0%, 7%) |
| *Muscle / joint pain* | 4 (1%)<br>(0%, 4%) | 3 (2%)<br>(0%, 7%) | 0 (0%)<br>(0%, 17%) | 0 (0%)<br>(0%, 16%) | 0 (0%)<br>(0%, 7%) |
| *Cognitive dysfunction* | 1 (<1%)<br>(0%, 2%) | 1 (1%)<br>(0%, 4%) | 0 (0%)<br>(0%, 17%) | 0 (0%)<br>(0%, 16%) | 0 (0%)<br>(0%, 7%) |
| *Visual disturbances* | 5 (2%)<br>(1%, 4%) | 3 (2%)<br>(0%, 7%) | 0 (0%)<br>(0%, 17%) | 1 (5%)<br>(0%, 24%) | 0 (0%)<br>(0%, 7%) |

\* By definition, all cases of PoTS were symptomatic during the Lean Test
† All patients meeting haemodynamic criteria for OH included including those with and without symptoms during Lean Test

#### Orthostatic Hypotension

21 (8%, 5% to 11%) of long covid participants met the criteria for OH, of which 11 (4%) had symptoms during the test and 10 (4%) were asymptomatic.

#### Orthostatic Tachycardia/PoTS

During the NLT, 28 (10%, 7% to 14%) participants experienced OT with no drop in blood pressure (data not shown), with 20 (7%, 4% to 11%) experiencing acute symptoms (dizziness, palpitations, chest pain or tremors) during the test, meeting the criteria for PoTS (Table 3). As shown in Figure 1, in patients who went on to meet the criteria for PoTS, the marked increase in mean heart rates was apparent shortly after standing, with a more gradual increase thereafter. By contrast, patients with negative test results initially increased by just over 10 bpm on average and remained stable for the remainder of the test. long covid patients with negative NLT results had mean heart rate increases similar to healthy volunteers with negative test results.

**Figure 1.**
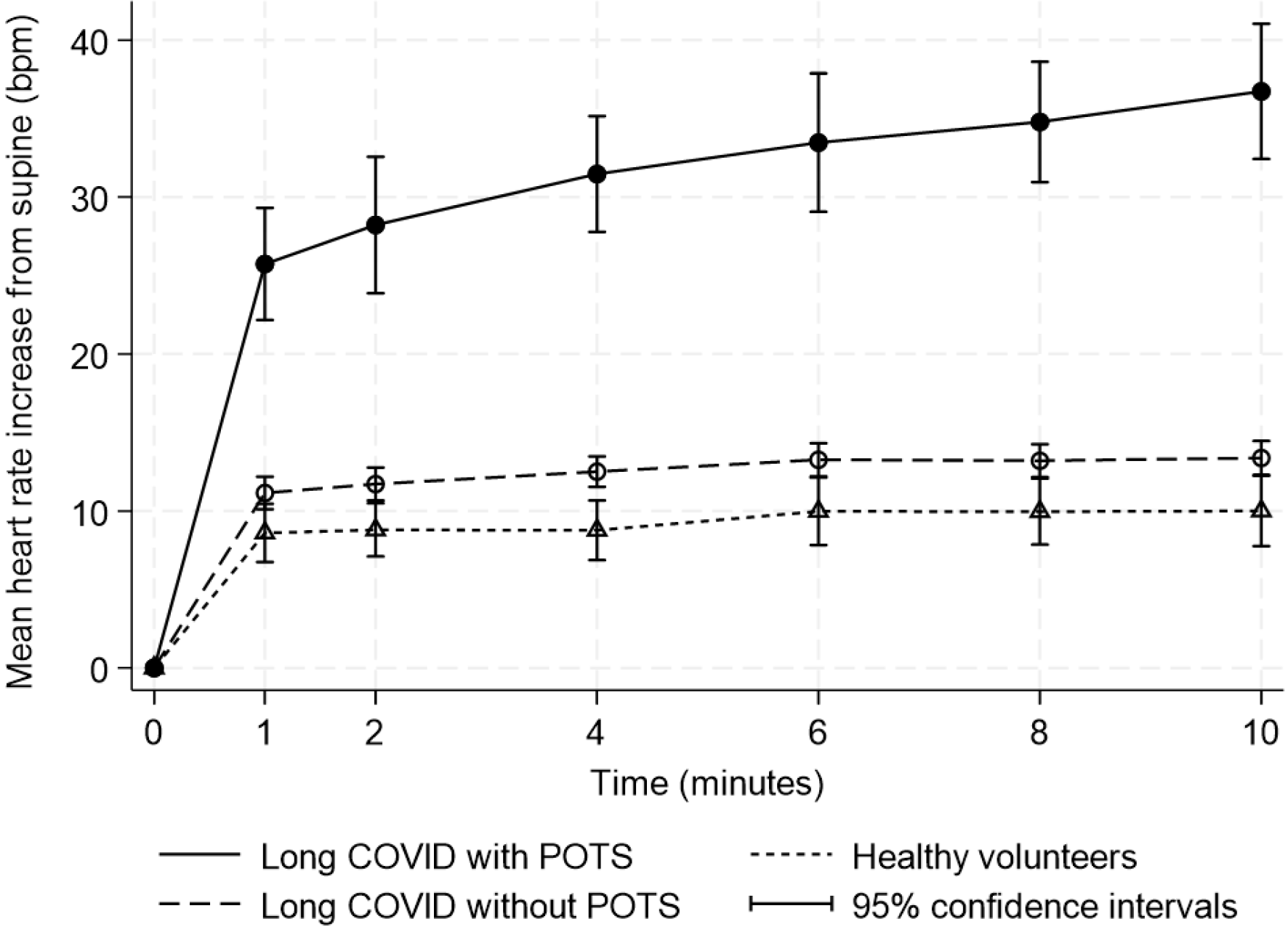
Mean heart rate increases for Long Covid patients who met the criteria for PoTS during the Lean Test, Long Covid patients who did not meet the criteria for PoTS during the Lean Test, compared to healthy volunteers who did not meet the criteria for PoTS

### Association between patient characteristics and prevalence of OI, OH, and PoTS

Symptoms experienced during the NLT are shown in Table 4. Associations between long covid patient characteristics, pre-existing comorbidities, and prior symptoms with PoTS and with OH are presented in Supplementary Tables 2 and 3 respectively. Those patients meeting the criteria for PoTS were younger and more likely to be female, report a higher prevalence of mental health conditions such as anxiety or depression, and more likely to have reported a history of typical OI symptoms compared to those with OH. Conversely, patients with OH were older with more physical health comorbidities.

### Association between prior history of OI and OH or PoTS

Whilst long covid patients who reported a prior history suggestive of OI, were more likely to meet the criteria for PoTS (RR=2.6, 1.0 to 6.7, p=0.03), 6 of the 20 had no history of OI symptoms (Table 3). By contrast, only a third of those who met the criteria for OH, 7 out of the 21, reported a history suggestive of OI. The occurrence of OH in those who did not report a history suggestive of OI was the same (10%) as that seen in the asymptomatic healthy volunteers (Table 3).

### Sensitivity to relaxing thresholds for diagnosing OH and PoTS

When the criterion for the increased heart rate to be sustained was relaxed, from two consecutive to one reading, this resulted in increasing the numbers of patients meeting the criteria for PoTS (from 7% to 11%) with no increase noted in healthy volunteers (Table 5). Increases were similar for those with and without a prior history of OI symptoms. In addition, decreasing the threshold from ≥30 bpm to ≥20 bpm resulted in approximately twice as many long covid patients meeting the criteria for PoTS. However, the increase in numbers that would meet the criteria was similar in patients with and without previous history of OI (Table 5). If the lower threshold was used, three (6%) healthy volunteers would have met the relaxed criteria for OT (data not shown), but still none would have met the criteria for PoTS because all healthy volunteers were asymptomatic during the test (Tables 4 and 5).

**Table 5.** Proportion of participants meeting conventional criteria for PoTS based on increased heart rate sustained over two consecutive readings ≥30 bpm, relaxed criteria based on a single reading ≥30 bpm, and relaxed criteria based on two consecutive readings ≥20 bpm.

|  | long covid patients (n=277) |  | All long covid patients (n=277) | Healthy volunteers (n=50) |
| --- | --- | --- | --- | --- |
|  | History of OI symptoms (n=130) | No history of OI symptoms (n=147) |  |  |
| Symptomatic during Lean Test with **two** consecutive heart rate readings $\geq 30$ bpm above supine (%) (95% CI) | 14 (11%)<br>(6%, 17%) | 6 (4%)<br>(2%, 9%) | 20 (7%)<br>(4%, 11%) | 0 (0%)<br>(0%, 7%) |
| Symptomatic during Lean Test with **single** heart rate reading $\geq 30$ bpm above supine (%) (95% CI) | 20 (15%)<br>(10%, 23%) | 11 (7%)<br>(4%, 13%) | 31 (11%)<br>(8%, 16%) | 0 (0%)<br>(0%, 7%) |
| Symptomatic during Lean Test with two consecutive heart rate readings **$\geq 20$ bpm** above supine (%) (95% CI) | 33 (25%)<br>(18%, 34%) | 17 (12%)<br>(7%, 18%) | 50 (18%)<br>(14%, 23%) | 0 (0%)<br>(0%, 7%) |

### Additional findings

Further analyses, reported in the Supplementary Table 4, include a difference in pulse pressure (PP) between long covid and healthy volunteers. While PP is not a routine clinical measurement, 122 (44%) of long covid patients were found to have a narrow PP (difference between diastolic and systolic, less than <25 of systolic blood pressure) compared to 10 (20%) of healthy volunteers. A narrow PP was seen more frequently, 71 out of 122 (58%), in long covid patients who reported acute symptoms during the NLT. There occurrence of a narrow PP was not associated with a history of OI, with 48% of long covid patients reporting a history suggestive of OI and 52% did not (data not shown).

## Discussion

These findings strongly confirm that OI, OH, and PoTS are all associated with a diagnosis of long covid, whilst OI, OH, and PoTS are rare in healthy volunteers. Whilst half of long covid clinic patients have symptoms of OI, not all reach the NLT threshold for either OH or PoTS. PoTS was more common in younger female patients, whilst OH was more common in older patients with comorbidities. Relaxing the criteria for sustained increase in heart rate would categorise more long covid patients with PoTS without resulting in healthy asymptomatic people being misclassified.

Previous studies of the prevalence of OI, OH, OT, and PoTS in long covid patients have been on small or unrepresentative samples,(24) with elevated rates of PoTS over 70% in a select cohort.(5,29) However, our estimates are consistent with those in comparable populations of 2-14%.(20,30,32) Whilst estimates of OH in long covid vary from 4-28%,(24,30) our results are comparable to similarly-aged cohorts.(24,30) Some differences in estimated prevalence can be explained by different lean test protocols, e.g. restricted diets,(34) which are not practical for routine clinic assessments. We applied commonly used criteria for PoTS, but implementation varies internationally. Others have previously applied a single elevated HR measurement,(9,32,34) an average of two,(37) the last three,(18) or two consecutive readings(6,30) These different thresholds have not previously been explored within the same cohort.

Acute symptoms during the NLT have been reported in 33-66% of patients with long covid population.(30,32) We saw similarly high prevalence of 53% (114 patients) with acute symptoms during the NLT which were not explained by haemodynamic measurements. This lack of association between symptoms and a positive NLT is consistent with previous findings.(20,30,32) Whilst most patients were able to complete a full 10 minutes of standing, 17% (46 patients) terminated the test early because of excessive symptoms, less than our previous report,(30) but higher than others have found.(32) Dizziness (65%, n=13) was the most frequent acute NLT symptom of PoTS, consistent with previous work,(5,32) but also tingling (30%, n=6), headaches (25%, n=5), breathlessness (15%, n=3) and chest discomfort (15%, n=3), contrasting with symptoms in people with OH of dizziness (25%, n=5), fatigue (14%, n=3), clamminess (14%, n=3) and tingling (10%, n=2).

Narrow Pulse Pressure (38) has previously been reported as low in long covid patients during a NLT,(34) consistent with our findings, supporting the role of dysautonomia and possible cerebral hypoperfusion (34) and providing an area for further research.

Potential limitations include that all patients were recruited from specialist long covid clinics, so prevalences may not be applicable in the community setting. We were unable to establish whether dysautonomia and OI are related to the severity of long covid, though previous work suggests this association is unlikely to be strong.(30) Our healthy volunteer comparator group was not individually matched but well-matched in overall age-sex distribution. Furthermore, our study was limited to UK patients and did adequately reflect a range of ethnic groups.

This is the largest study of prevalence of PoTS and OH in long covid to date, conducted across eight long covid clinics serving different populations, following standardised test administration, and including a healthy volunteer comparator group, allowing us to estimate the prevalence of PoTS and OH in long covid clinic patients. Our study also demonstrates the feasibility of real-world NLT testing across various clinical settings including self-administration at home. Assessing sensitivity to relaxing the criteria for diagnosis is a further strength of the work.

Our findings suggest the NLT is tolerated by most patients and should be considered as part of a holistic assessment to explore OI objectively in all patients with long covid rather than limiting it to those with typical symptoms of OI. Existing criteria for PoTS do not explain all symptoms and could be relaxed for those with at least one elevated HR rise >30 bpm with acute symptoms during the test or those with a sustained HR rise >20 bpm, or using modified versions of the test that allow capturing the triggers of OI such as exertion or food.(39)

We have demonstrated the value of the NLT in long covid as a safe and simple field test for OI, which can be performed either in a clinic setting or self-administered at home. Earlier detection of OI and dysautonomia should result in commencement of conservative measures and, if required, pharmacological management (4). Symptom trajectories and long-term outcomes of OI in long covid should be an area for future research (40). Health professionals supporting people living with long covid should be trained in the assessment, interpretation and management of OI and dysautonomia with clear pathways to further management.(41).

## Supporting information

Supplementary material - Lean test protocol

## Data Availability

All data produced in the present study are available upon reasonable request to the authors.

## Acknowledgments

We would like to acknowledge and thank the participants for their time and contribution.

