## Supplementary material - Lean test protocol for "Prevalence of orthostatic intolerance in Long Covid clinic patients: A multicentre observational study"

### Prevalence study of tachycardia on standing in long covid clinics

#### The quality improvement challenge

We don't know what best practice is in relation to postural orthostatic tachycardia syndrome (POTS, or fast heart rate on standing) in long covid clinics. Some people believe asymptomatic POTS is common in people with long covid (i.e. they claim we're missing it in clinic). Others say that clinically significant POTS is rare and almost always accompanied by symptoms such as dizziness or palpitations on standing. Some argue that since there's no treatment for POTS and tests may be falsely positive, there's no point looking for it. Some say testing for POTS leads to over-medicalisation.

We need to answer the question "what proportion of patients referred to long covid clinics have a significant increase in their heart rate on standing?". We think this may differ in different clinics. So this preliminary exercise is intended to **describe the clinic population**. What we're looking for is a statement such as 'In clinic X, 10% of patients meet the criteria for POTS whereas in clinic Y, only 2% do".

#### Who should be tested?

To compare across clinics, we need to all measure the same thing in the same way. **All patients should have the test**, since the point of this preliminary exercise is to get an accurate estimate for the **clinic**. Patients who have low probability of a positive test need to be included as well as those suspected of POTS. Anyone with a **clinical contra-indication** to the test (e.g. unable to stand safely) should be excluded. The full list of formal exclusion criteria to testing for POTS is available on request but clinical judgement is a good guide.

#### Testing for tachycardia on standing

Ask the patient to **lie down for 5 minutes**, then **measure their pulse and BP twice** (if the two readings are not similar, retake every 5 minutes until two consecutive readings are relatively close). The patient should then **stand up**—either without leaning on anything ('active stand test'), or leaning with just their shoulder blades against a wall ('lean test'). When they're comfortable, **measure pulse and BP every 2 minutes.**

| |
|---|
| Date of examination |
| Patient identifier (e.g. hospital number) |
| Symptoms suggestive of POTS?<br>e.g. dizziness or palpitations on standing<br>List all here |

| | Heart rate | Blood pressure | Symptoms during test |
|---|---|---|---|
| SUPINE READINGS | | | |
| 5 minutes (reading A) | | | |
| 5 minutes (reading B) | | | |
| STANDING READINGS | | | |
| 1 minute | | | |
| 2 minutes | | | |
| 4 minutes | | | |
| 6 minutes | | | |
| 8 minutes | | | |
| 10 minutes | | | |

When these tests are used in research studies, various additional data will be needed, but right now we just need the test readings and whether the patient has symptoms.

Pass completed forms to [ ] Any questions →

#### Test interpretation

Regardless of the test used, the interpretation of results to make a diagnosis is the same:

- **Orthostatic hypotension:** sustained reduction (for 10 minutes) of systolic blood pressure of at least 20 mmHg and/or diastolic blood pressure of at least 10 mmHg, or Systolic blood pressure fall >30 mmHg in hypertensive patients with supine systolic blood pressure > 160 mmHg, with upright position.
- **Orthostatic tachycardia or PoTS**: a sustained increase (for 10 minutes) in the patient's heart rate of at least 30 bpm (or 40 bpm or those patients aged 12-19 years old), a sustained increase of 40 bpm, or over 120 bpm in absence of orthostatic hypotension.
- **Inappropriate sinus tachycardia (IST)**: If the patient does not have orthostatic tachycardia but there is resting tachycardia, inappropriate sinus tachycardia (IST) should be considered.

If orthostatic intolerance is identified, differential diagnosis aimed at assessing underlying causes (e.g., anemia, endocrinological conditions, autoimmune condition) and appropriate treatment should be carried out.

See also the flow algorithm below taken from the *Canadian Cardiovascular Society Position Statement on Postural Orthostatic Tachycardia Syndrome (POTS) and Related Disorders of Chronic Orthostatic Intolerance*, Satish R. Raj, Juan C. Guzman, Paula Harvey, et al, Canadian Journal of Cardiology, Vol 36, Iss 3, 367-373, March 2020, [doi: 10/1016/j.cjca.2019.12.024]

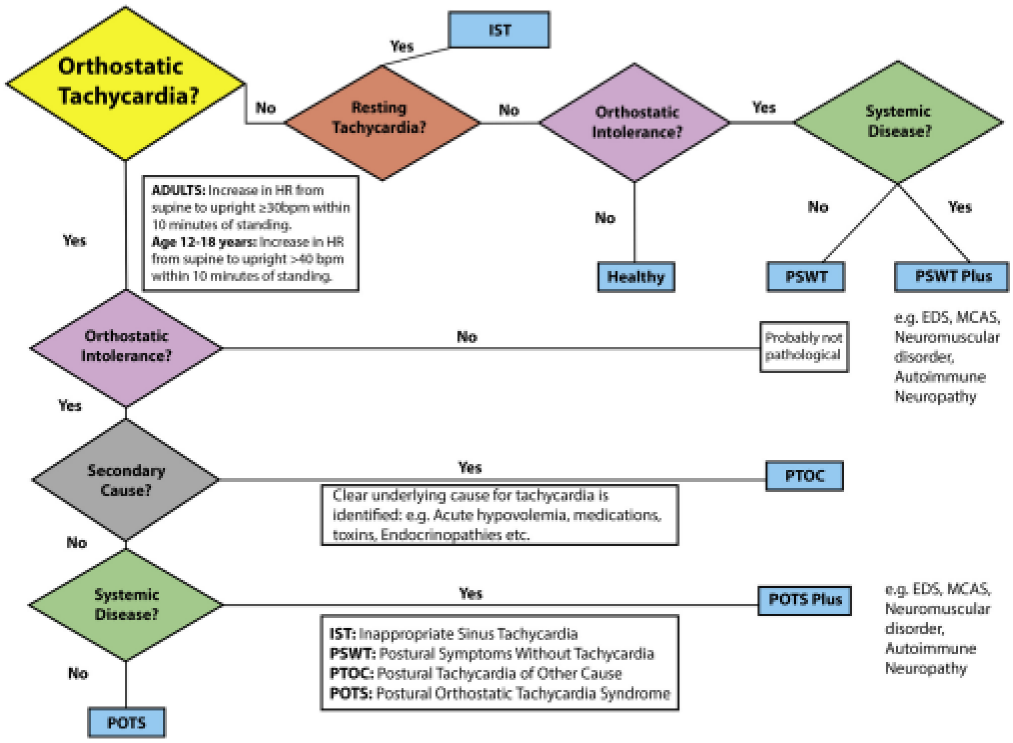

**Figure 2.** Postural orthostatic tachycardia syndrome (POTS) diagnostic criteria algorithm: a flow algorithm to help clinicians navigate the diagnosis of POTS and related disorders of orthostatic intolerance and orthostatic tachycardia. bpm, beats per minute; EDS, Ehlers-Danlos syndrome; HR, heart rate; IST, inappropriate sinus tachycardia; MCAS, mast cell activation syndrome; PSWT, postural symptoms without tachycardia.

#### Treatment options

Patients are often reassured when they receive an explanation of what is causing their symptoms. For those requiring intervention, consider non-pharmacological or conservative measures in the first instance.

If non-drug measures are not effective and symptoms are markedly affecting the patient's personal and professional life pharmacological options may be indicated, depending on the patient's medical history and clinical findings.

For a more detailed overview of treatment options see: *Orthostatic tachycardia after covid-19*, Ana B Espinosa-Gonzalez, Harsha Master, Nicholas Gall, Stephen Halpin, Natalie Rogers, Trish Greenhalgh ([BMJ 2023;380:e073488])
