## Supplementary tables for "Prevalence of orthostatic intolerance in Long Covid clinic patients: A multicentre observational study"

Supplemental material

Table 1. Acute test symptoms of dysautonomia on standing, presenting in Long COVID patients who could not complete the lean test because of excessive symptoms.

| | In Long COVID patients who could not complete lean test |
|---|---|
| Symptom | (n=46) |
| *Symptoms of orthostatic intolerance:* | |
| Dizziness / light-headedness | 23 (50%) |
| Palpitations | 5 (11%) |
| Chest pain / discomfort | 2 (4%) |
| Tremors | 1 (2%) |
| *Other symptoms of dysautonomia:* | |
| Tingling / numbness in skin | 11 (24%) |
| Fatigue | 11 (24%) |
| Sweating / clammy / hot | 5 (11%) |
| Breathlessness | 2 (4%) |
| Headache | 4 (9%) |
| Muscle / joint pain | 2 (4%) |
| Cognitive dysfunction | 1 (2%) |
| Visual disturbances | 1 (2%) |

Supplemental Table 2. Associations between patient characteristics, pre-existing comorbidities and prior symptoms with criteria for POTS, both unadjusted and adjusted for age and sex, in Long COVID patients.

| | POTS (n=20) | No POTS (n=257) | Unadjusted OR (95% CI) | Adjusted OR* (95% CI) |
|---|---|---|---|---|
| Patient characteristics (%): | | | | |
| Age ≥50 years | 5 (25%) | 139 (54%) | 0.3 (0.1, 0.8) | 0.3 (0.1, 0.8) |
| Female gender | 6 (30%) | 98 (38%) | 1.4 (0.5, 3.9) | 1.3 (0.5, 3.6) |
| Body mass index ≥30 kg/$m^2$ | 4 (27%) | 79 (41%) | 0.5 (0.2, 1.7) | 0.7 (0.2, 2.2) |
| Non-white ethnicity | 2 (10%) | 44 (18%) | 0.5 (0.1, 2.3) | 0.4 (0.1, 1.6) |
| Long COVID duration ≥12 months | 18 (90%) | 226 (88%) | 1.2 (0.3, 5.6) | 1.0 (0.2, 4.7) |
| Pre-existing comorbidities (%): | | | | |
| Allergies or autoimmune conditions | 4 (20%) | 40 (15%) | 1.4 (0.4, 4.3) | 1.6 (0.5, 5.2) |
| Other inflammatory conditions | 1 (5%) | 11 (4%) | 1.2 (0.1, 9.6) | 3.0 (0.3, 27.8) |
| Hypertension | 2 (10%) | 26 (10%) | 1.0 (0.2, 4.5) | 1.4 (0.3, 7.1) |
| Other heart conditions | 2 (10%) | 20 (8%) | 1.3 (0.3, 6.1) | 1.5 (0.3, 7.0) |
| Type 2 diabetes mellitus | 0 (0%) | 16 (6%) | - | - |
| Mental health condition | 6 (30%) | 37 (14%) | 2.5 (0.9, 7.1) | 2.2 (0.8, 6.3) |
| History suggestive of orthostatic intolerance (%): | | | | |
| Dizziness or light headedness | 14 (70%) | 91 (35%) | 4.3 (1.6, 11.5) | 3.9 (1.4, 10.8) |
| Palpitations | 7 (35%) | 63 (25%) | 1.7 (0.6, 4.3) | 1.4 (0.5, 3.7) |
| Chest pain or discomfort | 3 (15%) | 24 (9%) | 1.7 (0.5, 6.3) | 1.2 (0.3, 4.7) |
| Tremors | 2 (10%) | 9 (4%) | 3.1 (0.6, 15.2) | 4.2 (0.8, 22.9) |
| Any of dizziness, palpitations, chest pain, tremors | 14 (70%) | 116 (45%) | 2.8 (1.1, 7.6) | 2.3 (0.8, 6.3) |
| Other prior symptoms (%): | | | | |
| Fatigue | 3 (15%) | 27 (11%) | 1.5 (0.4, 5.5) | 1.5 (0.4, 5.7) |
| Aches or pains | 2 (10%) | 22 (9%) | 1.2 (0.3, 5.5) | 1.0 (0.2, 5.0) |
| Breathlessness | 2 (10%) | 14 (5%) | 1.9 (0.4, 9.2) | 2.3 (0.5, 11.2) |

* Odds ratios adjusted for age and sex. Age and sex only adjusted for each other.

Supplemental Table 3. Associations between patient characteristics, pre-existing comorbidities, and prior symptoms with criteria for orthostatic hypotension, both unadjusted and adjusted for age and sex, in Long COVID patients.

| | Orthostatic hypotension (n=21) | No Orthostatic hypotension (n=256) | Unadjusted OR (95% CI) | Adjusted OR* (95% CI) |
|---|---|---|---|---|
| Patient characteristics (%): | | | | |
| Age ≥50 years | 19 (90%) | 125 (49%) | 10.0 (2.3, 43.6) | 10.5 (2.4, 46.3) |
| Female gender | 8 (38%) | 96 (38%) | 1.0 (0.4, 2.4) | 1.4 (0.5, 3.6) |
| Body mass index ≥30 kg/m$^2$ | 5 (31%) | 78 (41%) | 0.7 (0.2, 2.0) | 0.4 (0.1, 1.4) |
| Non-white ethnicity | 2 (10%) | 44 (18%) | 0.5 (0.1, 2.3) | 0.8 (0.2, 3.9) |
| Long COVID duration ≥12 months | 19 (90%) | 225 (88%) | 1.3 (0.3, 5.9) | 1.7 (0.4, 8.1) |
| Pre-existing comorbidities (%): | | | | |
| Allergies or autoimmune conditions | 3 (14%) | 41 (16%) | 0.9 (0.2, 3.1) | 1.0 (0.3, 3.6) |
| Other inflammatory conditions | 1 (5%) | 11 (4%) | 1.1 (0.1, 9.1) | 0.7 (0.1, 6.1) |
| Hypertension | 4 (19%) | 24 (9%) | 2.3 (0.7, 7.3) | 1.9 (0.6, 6.4) |
| Other heart conditions | 1 (5%) | 21 (8%) | 0.6 (0.1, 4.4) | 0.6 (0.1, 4.8) |
| Type 2 diabetes mellitus | 4 (19%) | 12 (5%) | 4.8 (1.4, 16.4) | 2.6 (0.7, 9.5) |
| Mental health condition | 2 (10%) | 41 (16%) | 0.6 (0.1, 2.5) | 0.7 (0.1, 3.2) |
| History suggestive of orthostatic intolerance (%): | | | | |
| Dizziness or light headedness | 7 (33%) | 98 (38%) | 0.8 (0.3, 2.1) | 1.1 (0.4, 3.1) |
| Palpitations | 5 (24%) | 65 (25%) | 0.9 (0.3, 2.6) | 1.3 (0.4, 4.0) |
| Chest pain or discomfort | 0 (0%) | 27 (11%) | - | - |
| Tremors | 0 (0%) | 11 (4%) | - | - |
| Any of dizziness, palpitations, chest pain, tremors | 7 (33%) | 123 (48%) | 0.5 (0.2, 1.4) | 0.8 (0.3, 2.2) |
| Other prior symptoms (%): | | | | |
| Fatigue | 6 (29%) | 24 (9%) | 3.9 (1.4, 10.9) | 4.3 (1.4, 13.2) |
| Aches or pains | 1 (5%) | 23 (9%) | 0.5 (0.1, 3.9) | 0.6 (0.1, 5.1) |
| Breathlessness | 1 (5%) | 15 (6%) | 0.8 (0.1, 6.4) | 1.2 (0.1, 10.1) |

* Odds ratios adjusted for age and sex. Age and sex only adjusted for each other.

Supplemental Table 4. – Numbers and percentages of participants presenting during the lean test with sustained increased heart rate, orthostatic hypotension, or narrow pulse pressure, with and without acute symptoms during the lean test.

| | Long COVID patients (n=277) | | | | Healthy controls (n=64) | | | |
|---|---|---|---|---|---|---|---|---|
| | History of Long COVID symptoms suggestive of orthostatic intolerance § | | | | History of Long COVID symptoms suggestive of orthostatic intolerance § | | | |
| | No (n=147) | | Yes (n=130) | | No (n=50) | | Yes (n=0) | |
| Lean test measures | n (%) | (95% CI) | n (%) | (95% CI) | n (%) | (95% CI) | n (%) | (95% CI) |
| **Orthostatic tachycardia** | | | | | | | | |
| (Increased heart rate* with no drop in blood pressure†) | | | | | | | | |
| *With no acute symptoms during lean test* | 5 (3%) | (1, 8%) | 3 (2%) | (0, 7%) | 0 (0%) | (0, 7%) | - | - |
| *With acute symptoms during lean test (POTS)*‡ | 6 (4%) | (2, 9%) | 14 (11%) | (6, 17%) | 0 (0%) | (0, 7%) | - | - |
| **Orthostatic hypotension** | | | | | | | | |
| (Fall in blood pressure†) | | | | | | | | |
| *With no acute symptoms during lean test* | 6 (4%) | (2, 9%) | 4 (3%) | (0, 8%) | 5 (10%) | (3, 22%) | - | - |
| *With acute symptoms during lean test*‡ | 8 (5%) | (2, 10%) | 3 (2%) | (0, 7%) | 0 (0%) | (0, 7%) | - | - |
| **Narrow pulse pressure** | | | | | | | | |
| (Pulse pressure <25% of systolic blood pressure) | | | | | | | | |
| *With no acute symptoms during lean test* | 27 (18%) | (12, 26%) | 24 (18%) | (12, 26%) | 10 (20%) | (10, 34%) | - | - |
| *With acute symptoms during lean test*‡ | 31 (21%) | (15, 29%) | 40 (31%) | (23, 39%) | 0 (0%) | (0, 7%) | - | - |

* Heart rate increased by ≥30bpm (age 18+) or ≥40 bpm (age <18) sustained over two consecutive time points

† Fall in systolic blood pressure ≥20mmHg or diastolic blood pressure ≥10mmHg within first four minutes

‡ Symptoms of dysautonomia included dizziness, palpitations, chest pain or tremors, fatigue, muscle or joint pain, cognitive dysfunction, headache, visual disturbances, breathlessness, tingling or numbness in the skin, sweating or clamminess.

§ History of Long COVID symptoms suggestive of orthostatic intolerance prior to the lean test was taken as dizziness, palpitations, chest pain or tremors
